# The first 100 days of SARS-CoV-2 control in Vietnam

**DOI:** 10.1101/2020.05.12.20099242

**Authors:** Pham Quang Thai, Maia A Rabaa, Duong Huy Luong, Dang Quang Tan, Tran Dai Quang, Ha-Linh Quach, Ngoc-Anh Hoang Thi, Phung Cong Dinh, Ngu Duy Nghia, Tran Anh Tu, La Ngoc Quang, Tran My Phuc, Vinh Chau, Nguyen Cong Khanh, Dang Duc Anh, Tran Nhu Duong, Guy Thwaites, H Rogier van Doorn, Marc Choisy, OUCRU COVID-19 Research Group

## Abstract

**Background:** One hundred days after SARS-CoV-2 was first reported in Vietnam on January 23^rd^, 270 cases have been confirmed, with no deaths. We describe the control measures used by the Government and their relationship with imported and domestically-acquired case numbers, with the aim of identifying the measures associated with successful SARS-CoV-2 control.

**Methods:** Clinical and demographic data on the first 270 SARS-CoV-2 infected cases and the timing and nature of Government control measures, including numbers of tests and quarantined individuals, were captured by Vietnam’s National Steering Committee for COVID-19 response. Apple and Google mobility data provided proxies for population movement. Serial intervals were calculated from 33 infector-infectee pairs and used to estimate the proportion of pre-symptomatic transmission events and time-varying reproduction numbers.

**Findings:** After the first confirmed case on January 23^rd^, the Vietnamese Government initiated mass communications measures, case-contact tracing, mandatory 14-day quarantine, school and university closures, and progressive flight restrictions. A national lockdown was implemented between April 1^st^ and 22^nd^. Around 200 000 people were quarantined and 266 122 RT-PCR tests conducted. Population mobility decreased progressively before lockdown. 60% (163/270) of cases were imported; 43% (89/208) of resolved infections remained asymptomatic for the duration of infection. 21 developed severe disease, with no deaths. The serial interval was 3.24 days, and 27.5% (95% confidence interval, 15.7%-40.0%) of transmissions occurred pre-symptomatically. Limited transmission amounted to a maximum reproduction number of 1.15 (95% confidence interval, 0.37-2.36). No community transmission has been detected since April 15^th^.

**Interpretation:** Vietnam has controlled SARS-CoV-2 spread through the early introduction of mass communication, meticulous contact-tracing with strict quarantine, and international travel restrictions. The value of these interventions is supported by the high proportion of asymptomatic and imported cases, and evidence for substantial pre-symptomatic transmission.

**Funding:** The Vietnam Ministry of Health and Wellcome Trust, UK.

**Research in context:** *Evidence before this study:* Vietnam was one of the first countries outside of China to detect imported and human-to-human transmitted SARS-CoV-2 within its borders. Yet, as of May 1^st^, a total of only 270 cases have been confirmed, no deaths have occurred, and no community transmission has been detected since April 15^th^ despite intensive screening, tracing and testing. We did a PubMed database search to identify studies investigating COVID-19 response in Vietnam using the terms “Vietnam”, “COVID-19”, and “SARS-CoV-2”. All relevant articles were evaluated. Studies describe cases of COVID-19 and their management, aspects of the government response from newspapers and online government sources, but there are no previous reports using national data to describe and investigate the national epidemic and the impact of control measures cases over time.

*Added value of this study:* We used data from the National Steering Committee for COVID-19 response to give a comprehensive account of the first 100 days of the SARS-CoV-2 epidemic in Vietnam, including case numbers and their symptomatology, the estimated reproductive number by week, and their relation to the multiple control measures instituted by the Vietnam Government over time. We show two distinctive features of Vietnam’s response. First, the Government took rapid actions to restrict international flights, closed schools and universities, and instituted meticulous case-contact tracing and quarantining from late January, well before these measures were advised by WHO. Second, they placed mass communication, education, and the identification, serial testing, and 14-day quarantine of all direct contacts of cases, regardless of symptom development, at the heart of the response. The value of strict contact-tracing and quarantine is supported by the high proportion of asymptomatic cases (43%) and imported cases (60%), and evidence for substantial pre-symptomatic transmission.

*Implications of all the available evidence:* Vietnam has had remarkable success in controlling the emergence of SARS-CoV-2. Our report provides a complete picture of the control of SARS-CoV-2 in Vietnam, with lessons for other Governments seeking to extend national SARS-CoV-2 control or prevent future epidemics. Our findings shows the importance of acting early, before the virus becomes established in the community, and before the case numbers overwhelm systems of case-contact tracing and mass quarantine. They also demonstrate the value of effective mass communication in rapidly educating the public in infection prevention measures and providing real-time information on the state of the epidemic.

## Introduction

The severe acute respiratory syndrome coronavirus 2 (SARS-CoV-2) emerged in Wuhan city, Hubei Province, China, in late 2019^1^. On January 30^th^, the WHO declared the outbreak a ‘Public Health Emergency of International Concern’, and on March 11^th^ a global pandemic. By May 1^st^ 2020, the virus had infected more than 3 million people and killed over 200 000. The numbers of cases and deaths continue to rise in many countries worldwide and the virus now represents the greatest acute infectious threat to humankind since the influenza pandemic of 1918.

SARS-CoV-2 is antigenically different from known human and zoonotic coronaviruses and there is no known pre-existing population immunity^2^. It is highly transmissible through respiratory secretions expelled from an infected person, with a basic reproduction number (R_0_) estimated between 2 and 3 in the absence of control measures^3-6^. Many infections are asymptomatic^7^, while others lead to symptoms of coronavirus disease (COVID-19) of varying severity^5^. Analyses of serial intervals suggest that contagiousness can occur both before and after the onset of symptoms as well as in those who never develop symptoms^8^. The subsequent exponential rise in infections has threatened to overwhelm even the world’s best developed health systems and cause major loss of life. Methods to control the virus and reduce the impact of COVID-19 have thus become a global priority.

The preparedness, timing, and nature of the response to SARS-CoV-2 have varied substantially between countries. Many affected countries have resorted to extreme social distancing measures through so-called ‘lockdowns’, where populations isolate themselves within their homes, reducing all but essential contact with others. As first observed in Hubei Province in China, and subsequently in other countries, these measures slow transmission and reduce disease incidence, but at significant social and economic cost^9-11^. However, ‘lockdowns’ represent a combination of potentially independent interventions (for example, closing schools and universities, suspending public transport, banning public gatherings, closing non-essential businesses), the effects of which in isolation are uncertain. Determining their relative contributions to SARS-CoV-2 control is critical to understanding how they might be safely and incrementally lifted, or partially reinstated. Such information may be acquired from studying the measures employed by countries that have so far controlled the virus.

Vietnam is a low-middle income country that shares borders with China, The Lao People’s Democratic Republic, and Cambodia. It is the 15^th^ most populous country on earth, with 97.3 million people, and it was one of the first countries affected by SARS-CoV-2, recording its first case on January 23^rd^ 2020. Yet, as of May 1^st^, 270 cases have been confirmed, with no deaths^12^. In this report we describe the first 100 days of SARS-CoV-2 control in Vietnam, including the timing and types of interventions and their impacts on imported and domestically-transmitted case numbers. Our aim was to identify the measures most closely associated with successful SARS-CoV-2 control.

## Methods

Clinical, epidemiological and policy data were provided by Vietnam’s National Steering Committee for COVID-19 response. Data from 270 SARS-CoV-2-confirmed cases to May 1^st^ 2020 included their age, gender, nationality, dates of symptom onset (if any), entry to the country and quarantine (if any), hospital admission and discharge, and the results of all RT-PCR tests. Imported cases were distinguished from those acquired domestically, with information on quarantine at or after entry to the country. Imported cases were denoted G0; and among domestically-acquired infections, those acquired directly from G0 cases were denoted as G1, others were denoted G2+.

Intervention data consisted of daily time-series of the numbers in quarantine and RT-PCR tests performed. Daily reports from the Ministry of Health and Vietnam’s National Steering Committee for COVID-19 response listed key milestones in national SARS-CoV-2 control measures. Apple mobility data^13^ and Google community mobility data^14^ provided proxies of population movements, with additional information provided in the Supplementary Appendix.

Serial intervals were calculated from infector-infectee pairs and fitted to a normal distribution by maximum likelihood^8^. The estimated distribution parameters (mean and standard deviation, together with their confidence intervals and variance-covariance matrix) were used to estimate the proportion of pre-symptomatic transmissions and three time-varying reproduction numbers^15^: between G0 and G1 (step 1), between G1 and G2+ (step 2), and between G0, G1, and G2+ combined (step 1 and 2 combined). Further details are provided in the Supplementary Appendix.

We used a logistic regression to investigate the link between the proportion of asymptomatic infections and age, gender, nationality (Vietnamese versus non-Vietnamese), and imported versus domestically-acquired infection. We used a gamma regression to investigate the link between the duration of hospitalisation and the same variables listed above, plus symptomatic versus asymptomatic. To correct for potential confounding effects between the explanatory variables, we used Type-II likelihood ratio tests^16^. All analyses were done with R 4.0.0^17^ using the packages EpiEstim^18^ 2.2-1 and fitdistrplus^19^ 1.0-14. Further details of the analysis are provided in the Supplementary Appendix.

## Results

### Epidemic description and control measures

On January 10^th^, before the first case was confirmed in Vietnam, the Vietnam Government reinforced temperature and health status screening at border gates for passengers arriving from Wuhan, tracing and quarantining of suspected cases and their contacts, monitoring of suspected cases of respiratory infections in hospitals and the community, and initiated mass communication to the public on preventive measures (including hand washing, contact avoidance and mask wearing).

The epidemic timeline for Vietnam, including the numbers quarantined and hospitalised, tests performed, cases confirmed, population movements, and the timing and nature of major Government-led control measures are summarised in **Figure 1**. The control measures are also summarised in **Table 1** and in greater detail in **Table S1**. To date, two waves of transmission have occurred: the first began on January 23^rd^ and resulted in 16 cases (9 imported, 7 acquired in-country), and the second on March 6^th^, leading to 254 cases (154 imported, 100 acquired in-country).

**Figure 1.**
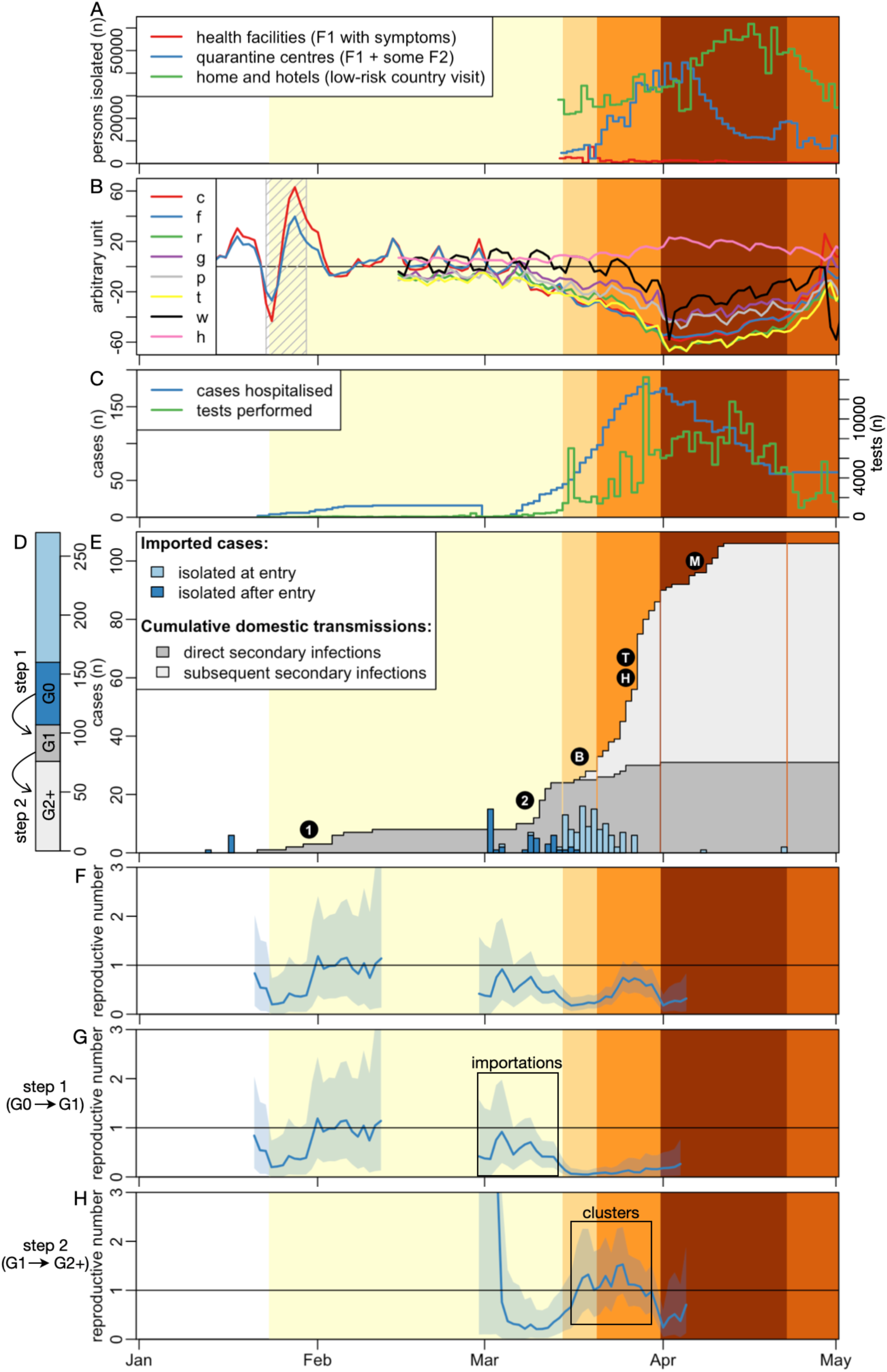
Timeline of SARS-CoV-2 emergence and response in Vietnam. The background colour reflects the intensity of the interventions taken by the Vietnam Government to control the COVID-19 epidemic, with darker shades indicating more intense disease control measures. The main events of these periods are described in detail in **Table 1**. **Panel A:** number of people in isolation by day. **Panel B:** relative indices of population movements: number of travellers by car (**c**), on foot (**f**) (both from Apple Mobility Data^13^), proxies of people in retail and recreation areas (**r**), in groceries stores and pharmacies (**g**), in parks (**p**), in bus transit stations (**t**), at work (**w**), and at home (**h**), all from Google Community Mobility Data^14^. The hashed area indicates the lunar New Year holiday (23-29^th^ January). Traditionally, the first half of the week is spent at home with close family, whereas the second half of the week is dedicated to visits of members of the extended family. **Panel C:** number of SARS-CoV-2 positive cases hospitalised and RT-PCR tests performed by day. **Panel D:** cumulative number of detected SARS-CoV-2 positive cases in Vietnam, differentiating imported cases (G0) and whether they were isolated at entry or later, and locally transmitted cases and whether they were in direct contact with imported cases (G1) or not (G2+)). **Panel E:** numbers of SARS-CoV-2 imported cases together with cumulative numbers of local transmissions. Circled characters indicate major internal transmission events: first introduction of SARS-CoV-2 virus in the country (**1:** 16 cases), second introduction (**2:** 15 cases), cluster of transmission in a Ho Chi Minh City bar (**B:** 19 cases), cluster of transmission in a large Hanoi hospital (**H:** 17 cases), community cluster of transmission linked to the Hanoi hospital through catering staff (**T:** 28 cases) and community cluster of transmission in Me Linh district in the north of Hanoi (**M:** 13 cases). **Panels F-H:** estimates of the reproduction number for the two epidemics. **G** focuses only on the first step of the chain of transmission between G0 and G1, whereas **H** focuses on all the other steps of the chain of transmission. **F** includes all detected cases. The shaded blue area shows the 95% confidence intervals.

**Table 1.**
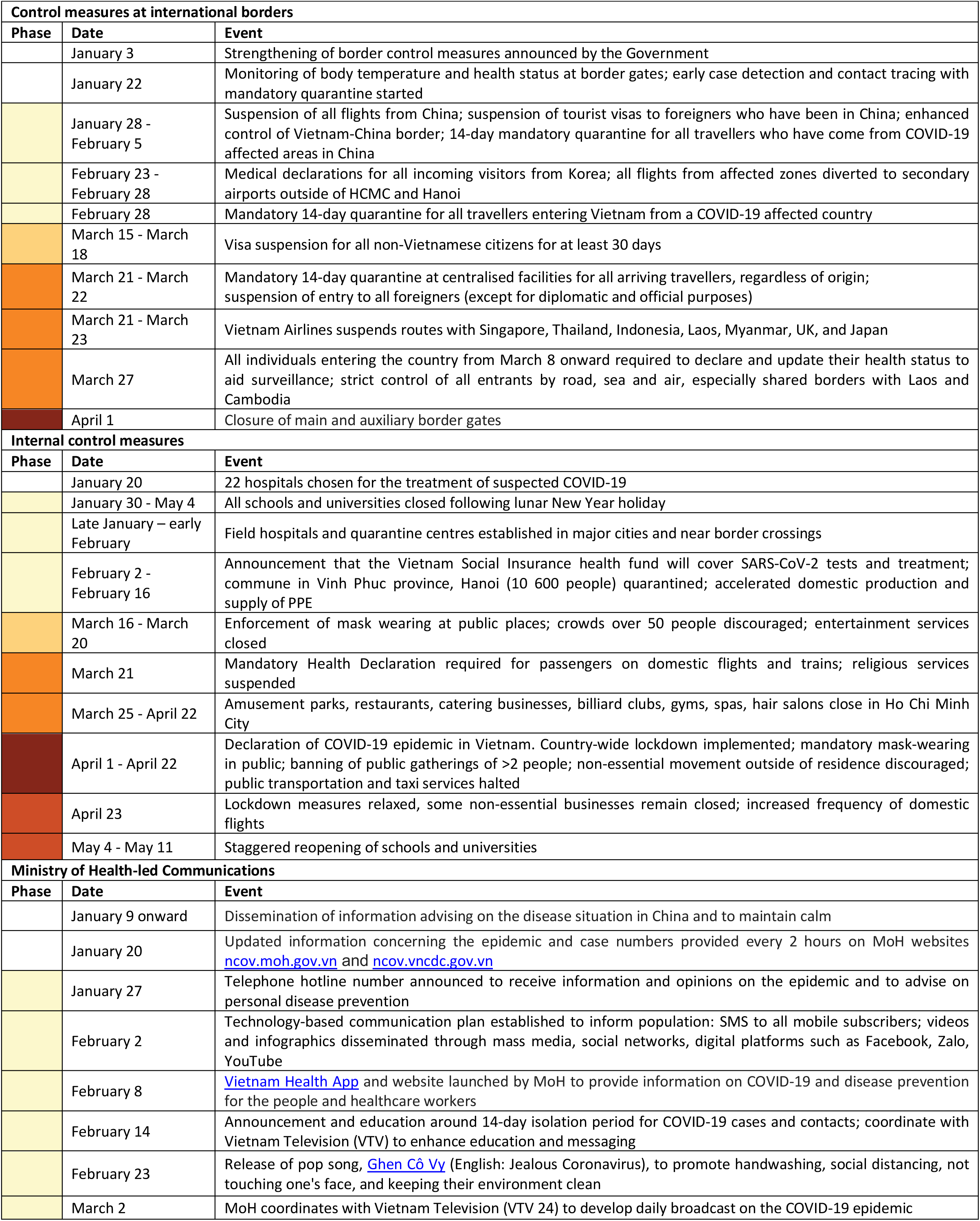
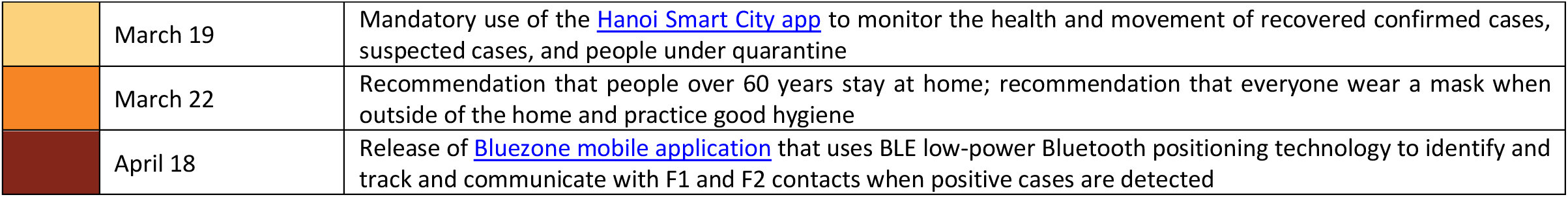
The timing and nature of major Vietnam Government-led control measures, including international border control, internal control, and Ministry of Health-led communications. (further details provided in **Table S1**). The colours shown in the phase column indicate the intensity of control measures taken over different periods (white, initial; light yellow, early; light orange, intermediate; orange, pre-epidemic; brown, epidemic/lockdown; dark orange, post-lockdown), and correspond to those used in **Figure 1** and **Table S1**.

| Control measures at international borders |  |  |
| --- | --- | --- |
| Phase | Date | Event |
|  | January 3 | Strengthening of border control measures announced by the Government |
|  | January 22 | Monitoring of body temperature and health status at border gates; early case detection and contact tracing with mandatory quarantine started |
|  | January 28 - February 5 | Suspension of all flights from China; suspension of tourist visas to foreigners who have been in China; enhanced control of Vietnam-China border; 14-day mandatory quarantine for all travellers who have come from COVID-19 affected areas in China |
|  | February 23 - February 28 | Medical declarations for all incoming visitors from Korea; all flights from affected zones diverted to secondary airports outside of HCMC and Hanoi |
|  | February 28 | Mandatory 14-day quarantine for all travellers entering Vietnam from a COVID-19 affected country |
|  | March 15 - March 18 | Visa suspension for all non-Vietnamese citizens for at least 30 days |
|  | March 21 - March 22 | Mandatory 14-day quarantine at centralised facilities for all arriving travellers, regardless of origin; suspension of entry to all foreigners (except for diplomatic and official purposes) |
|  | March 21 - March 23 | Vietnam Airlines suspends routes with Singapore, Thailand, Indonesia, Laos, Myanmar, UK, and Japan |
|  | March 27 | All individuals entering the country from March 8 onward required to declare and update their health status to aid surveillance; strict control of all entrants by road, sea and air, especially shared borders with Laos and Cambodia |
|  | April 1 | Closure of main and auxiliary border gates |
| Internal control measures |  |  |
| Phase | Date | Event |
|  | January 20 | 22 hospitals chosen for the treatment of suspected COVID-19 |
|  | January 30 - May 4 | All schools and universities closed following lunar New Year holiday |
|  | Late January – early February | Field hospitals and quarantine centres established in major cities and near border crossings |
|  | February 2 - February 16 | Announcement that the Vietnam Social Insurance health fund will cover SARS-CoV-2 tests and treatment; commune in Vinh Phuc province, Hanoi (10 600 people) quarantined; accelerated domestic production and supply of PPE |
|  | March 16 - March 20 | Enforcement of mask wearing at public places; crowds over 50 people discouraged; entertainment services closed |
|  | March 21 | Mandatory Health Declaration required for passengers on domestic flights and trains; religious services suspended |
|  | March 25 - April 22 | Amusement parks, restaurants, catering businesses, billiard clubs, gyms, spas, hair salons close in Ho Chi Minh City |
|  | April 1 - April 22 | Declaration of COVID-19 epidemic in Vietnam. Country-wide lockdown implemented; mandatory mask-wearing in public; banning of public gatherings of >2 people; non-essential movement outside of residence discouraged; public transportation and taxi services halted |
|  | April 23 | Lockdown measures relaxed, some non-essential businesses remain closed; increased frequency of domestic flights |
|  | May 4 - May 11 | Staggered reopening of schools and universities |
| Ministry of Health-led Communications |  |  |
| Phase | Date | Event |
|  | January 9 onward | Dissemination of information advising on the disease situation in China and to maintain calm |
|  | January 20 | Updated information concerning the epidemic and case numbers provided every 2 hours on MoH websites ncov.moh.gov.vn and ncov.vncdc.gov.vn |
|  | January 27 | Telephone hotline number announced to receive information and opinions on the epidemic and to advise on personal disease prevention |
|  | February 2 | Technology-based communication plan established to inform population: SMS to all mobile subscribers; videos and infographics disseminated through mass media, social networks, digital platforms such as Facebook, Zalo, YouTube |
|  | February 8 | Vietnam Health App and website launched by MoH to provide information on COVID-19 and disease prevention for the people and healthcare workers |
|  | February 14 | Announcement and education around 14-day isolation period for COVID-19 cases and contacts; coordinate with Vietnam Television (VTV) to enhance education and messaging |
|  | February 23 | Release of pop song, Ghen Cô Vy (English: Jealous Coronavirus), to promote handwashing, social distancing, not touching one's face, and keeping their environment clean |
|  | March 2 | MoH coordinates with Vietnam Television (VTV 24) to develop daily broadcast on the COVID-19 epidemic |

|  |  |
| --- | --- |
| March 19 | Mandatory use of the Hanoi Smart City app to monitor the health and movement of recovered confirmed cases, suspected cases, and people under quarantine |
| March 22 | Recommendation that people over 60 years stay at home; recommendation that everyone wear a mask when outside of the home and practice good hygiene |
| April 18 | Release of Bluezone mobile application that uses BLE low-power Bluetooth positioning technology to identify and track and communicate with F1 and F2 contacts when positive cases are detected |

The first confirmed cases of SARS-CoV-2 infection presented in Hanoi and Ho Chi Minh City during the lunar New Year holiday (23-29^th^ January). Cases were travellers from Wuhan city or their contacts, and were identified by the public health laboratory network using improvised molecular diagnostics, including agnostic sequencing, prior to implementation of the WHO-approved assays^20^. Amongst the cases were the first confirmed human-to-human transmissions outside of China^21^.

Entry of airline passengers into Vietnam from Wuhan city and elsewhere in China was monitored and progressively limited (**Table 1**), and cases and their contacts were quarantined for 14 days in Government facilities to prevent onward transmission. Schools and universities remained closed after the lunar New Year holiday, with staggered re-opening from May 4^th^ (closures lasted ~3 months). The National Steering Committee for COVID-19 response was established in late January, composed of 24 members from 23 ministries charged with coordinating the epidemic response. A hotline was set-up by the Ministry of Health on January 27^th^, a nationwide SMS push notification system was put in place through all mobile phone providers on February 3^rd^, and a mobile phone app for contact tracing and symptom reporting was launched on February 8^th^.

In early February, following the repatriation of a number of Vietnamese nationals from Wuhan city, a cluster of community transmitted infections was detected in two communes in Vinh Phuc province, bordering Hanoi, including a 3-month old infant^22,23^. On February 13^th^, these communes were quarantined for three weeks, with no additional cases detected in the country until March 6^th^ and the start of the second wave of infections in Hanoi.

This second wave began on March 6^th^ following diagnosis of the index case, who had arrived in Hanoi on March 2^nd^ from London after visiting Italy and the UK. Following the identification of this case, all passengers and crew who had been on the flight from London with the index case were quarantined in Government facilities for 14 days, as were all individuals in direct contact with the index or any subsequent cases. The immediate neighbourhood of the index case was sealed off, with active surveillance conducted to detect any new cases. These surveillance measures revealed SARS-CoV-2 infection in 12 others on the flight and 2 close contacts of the infected traveller after entering Vietnam.

Further cases occurred in the following two weeks, mostly in foreign and returning Vietnamese travellers from Europe and the USA, including multiple acquisitions in a Ho Chi Minh City bar on March 14^th^ (19 cases), a cluster among nursing (17 cases) and catering (28 cases) staff in a large Hanoi hospital, and a community cluster in Me Linh district (13 cases), in the north of Hanoi. Systematic layered testing and quarantine requirements were put in place for cases (F0) and their direct (F1) and indirect (F2-4) contacts. Cases were isolated in assigned hospitals until tested negative at least twice by RT-PCR. F1 and F2 contacts were quarantined for at least 14 days in dedicated facilities (health centres, hotels, military camps) with negative tests required before release. F3 and F4 contacts were asked to self-quarantine for 14 days. Until May 1^st^, around 70 000 have been quarantined in Government facilities and around 140,000 at home or in hotels. 266 122 RT-PCR-based SARS-CoV-2 tests were performed, with a ratio of around 1 positive person: 1 000 tests conducted.

After further measures to prevent entry of infected international travellers (**Table 1**), a nationwide lockdown was enforced on April 1^st^, including closure of all shops except gas stations, food stores, and pharmacies; suspension of public transport, including all taxis; and mandatory mask wearing in all public spaces. Mobility data show that population movement decreased substantially after the start of the second infection wave in early March, reaching a nadir in early April at the start of the lockdown (**Figure 1**). Movements increased slowly during the last week of the lockdown and more rapidly once the lockdown was partially lifted on April 23^nd^. On April 15^th^, the last case of wave two was identified; subsequent cases (n = 2) have been detected between April 15^th^ and May 1^st^ (time of writing) among international travellers quarantined on arrival.

### Characteristics of the cases

Sixty percent (163/270) of cases were imported (**Figure 2**); 110 were quarantined and diagnosed on entry, whereas 53 entered prior to the implementation of systematic quarantine measures and were identified in the community. Vietnamese nationals represented 134/163 (82.2%) of the imported cases and 89/107 (83.2%) of those acquired in-country. The median age of imported and domestically-acquired cases was 27 years (interquartile range (IQR) 21-42) and 41 years (IQR 28-49), and 81/163 (50.0%) and 69/107 (64.5%) of these were female, respectively.

**Figure 2.**
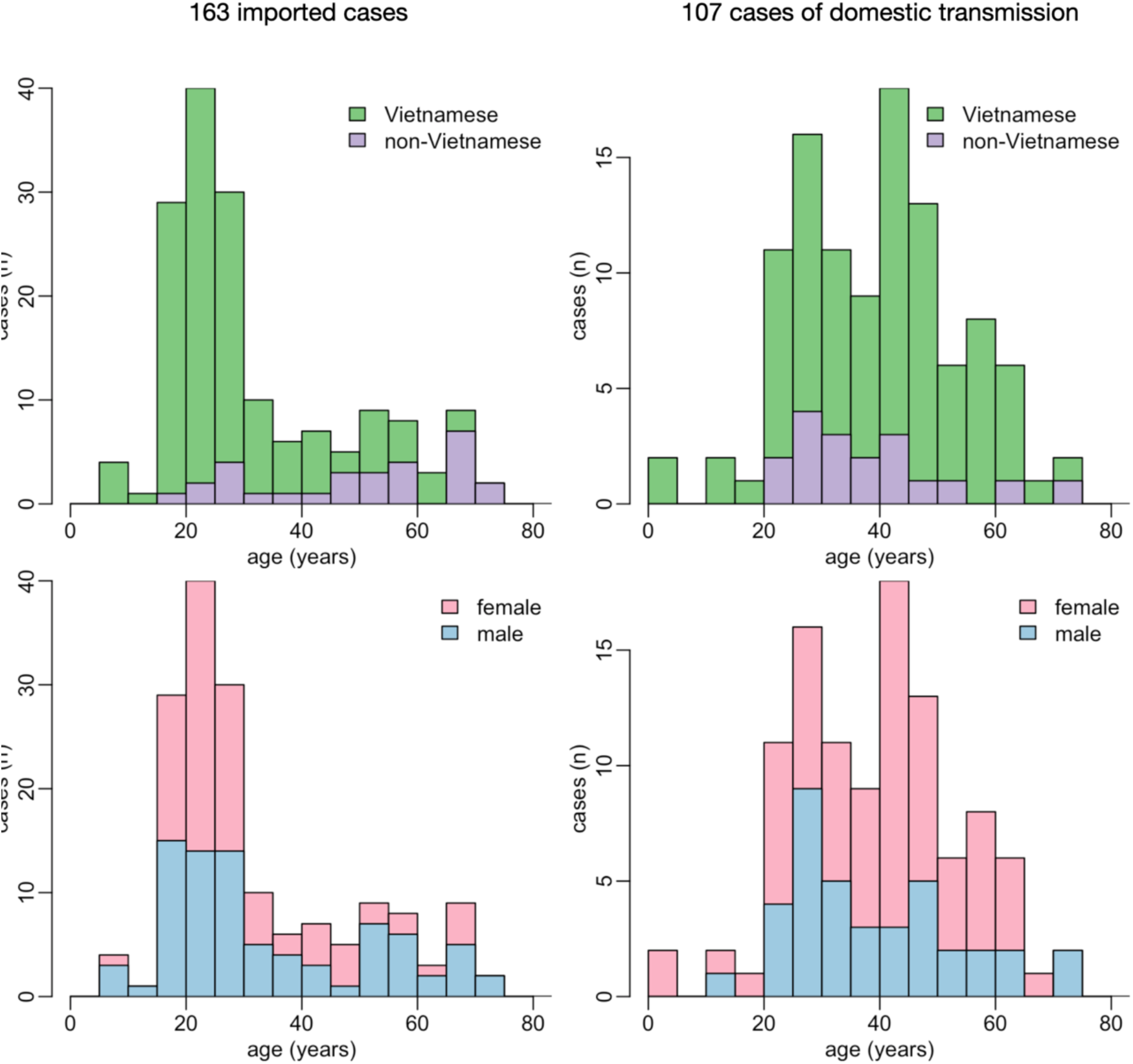
Demographics of the 270 SARS-CoV-2 positive cases in Vietnam. Age distribution for the 163 imported cases (left column) and the 107 cases of local transmission (right column), by nationality (top row) and gender (bottom row).

By May 1^st^, 208 patients were discharged and 62 remained hospitalised for treatment or isolation. Forty-three percent (89/208) of discharged cases never developed symptoms, and this was not significantly associated with age, gender, nationality, or origin of infection (imported or domestically-acquired). Symptoms developed after entry to the country in 73.9% (68/92) and in a Government quarantine facility in 33.9% (38/150) (**Figure 3**). The median age of symptomatic and asymptomatic cases was 30 (IQR 24-49) and 31 (IQR 23-45), respectively (**Figure 3**). Among the 150 with symptoms, 21 (14.0%) developed severe disease, of whom five required mechanical ventilation and two received extra-corporeal membrane oxygenation. No fatalities were recorded. The duration of hospitalisation was significantly shorter (p<0.0001) for asymptomatic (17 days, IQR 13-22) than for symptomatic cases (19 days, IQR 16-25). While gender, nationality, and origin of infection did not have any significant effect, the duration of hospitalisation of symptomatic cases increased with age (**Figure 3**).

**Figure 3.**
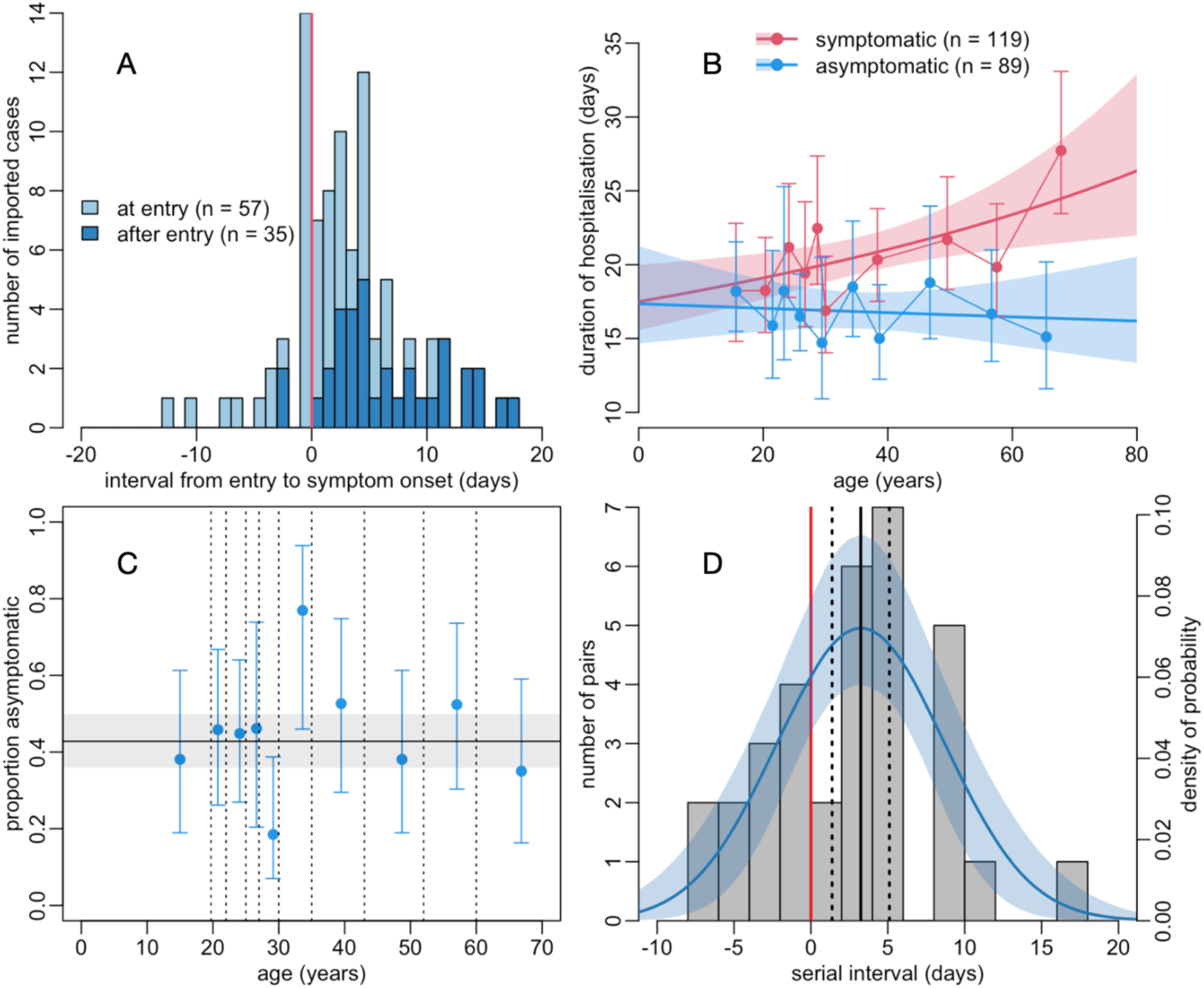
Asymptomatic and symptomatic SARS-CoV-2 infection in Vietnam. **Panel A:** distribution of the interval between entry into the country and the onset of symptoms for 92 symptomatic imported SARS-CoV-2 positive cases, differentiating those who were isolated at entry from those who were not. Symptoms occurred after entry on the right-hand side of the vertical red line. **Panel B:** duration of hospital stay of 208 discharged SARS-CoV-2 positive cases. Dots and error bars show mean and 95% confidence interval (assuming a gamma distribution) per decile of age, lines and shaded areas show gamma regression fits and their 95% confidence intervals. The corresponding gamma regression table is in **Table S2**. **Panel C:** relationship between age and the proportion asymptomatic among 208 discharged SARS-CoV-2 positive cases. Vertical dotted lines indicate deciles of the age distribution, with the proportion asymptomatic estimated within each of these deciles. Vertical error bars show 95% confidence intervals. The horizontal line and the grey area show the average across ages and its 95% confidence interval. The corresponding logistic regression table is in **Table S3**. **Panel D:** distribution of serial intervals for 33 infector-infectee pairs together with a normal distribution fitted to it. The shaded area shows the 95% confidence interval. The vertical black line shows the estimate of the mean serial interval, together with its 95% confidence interval (dashed vertical lines). The proportion of the distribution to the left of the red line is a proxy for the proportion of infections that occur before the onset of symptoms.

### Epidemiological parameters over time

From 33 infector-infectee pairs, the mean serial interval was estimated to be 3.24 days (95% confidence interval (CI), 1.38-5.10 days) with a standard deviation of the distribution of 5.46 days (95% CI, 4.14-6.78 days). An estimated 27.5% (95% CI, 15.7%-40-0%) of the distribution was below zero, suggesting these transmissions occurred prior to the onset of symptoms in the infector. From the (non-quarantined) imported cases (G0) and onward infected cases (G1 and G2+), we calculated the effective reproductive number R by date (**Figure 1**). Limited transmission amounted to a maximum R of 1.15 (95% CI, 0.37-2.36). R rarely exceeded 1 and a decrease of R is seen as more mitigating measures were implemented from the end of March before the nationwide lockdown. When analysing R from G0 to G1 (step 1) and from G1 to G2+ (step 2) separately, we found that R was drastically decreased for step 1 simultaneously with suspension of all international travel (March 18^th^), while for step 2 transmission continues with R slightly above 1 despite intense contact tracing and quarantine. Only during the nationwide lockdown R was reduced to less than 1 (Figure 1).

## Discussion

On January 23^rd^ 2020, Vietnam was one of the first countries to report SARS-CoV-2 infection and the first to report human-to-human transmission outside of China^21^. Yet 100 days later, it confirmed just 270 cases despite extensive testing, with no community transmission since April 15^th^. In the three weeks prior to May 1^st^, there were only two imported cases and no reported cases elsewhere in the country. The nature, timing, and success of the control measures introduced may have relevance to other countries seeking to control SARS-CoV-2 transmission.

Vietnam has experience in responding to emerging infectious diseases. In the last 20 years, it has confronted outbreaks of SARS^24^, avian and pandemic influenza^25,26^, hand-foot-and-mouth disease^27^, measles^28^, and dengue^29^. Its outbreak responses are coordinated by the Ministry of Health, a permanent national Public Health Emergency Operations Centre at the National Institute for Hygiene and Epidemiology, and through a network of provincial Centres for Disease Control and lower level preventive medicine centres^30^.

Two waves of SARS-CoV-2 infections have occurred over the last 100 days in Vietnam, with community transmission actively interrupted by rapid isolation and identification of primary and secondary cases and their contacts. To date, around 200 000 people have spent at least 14 days in quarantine. Amongst those quarantined, many were second degree contacts (F2); to our knowledge, no other country has implemented quarantine in this manner. 266 122 RT-PCR tests have been performed, primarily in those quarantined, giving the highest global ratio of tests conducted per positive person (~1000:1).

The majority of cases (60%) in Vietnam were imported among those arriving from COVID-19 affected countries; first from China and then from countries in Europe and the USA. Early introduction of airport screening, followed by quarantine of all arrivals and the eventual suspension of nearly all international flights prevented further introductions, allowing greater focus on the detection and prevention of domestic transmission. Consistent and pervasive Government communication of disease risk and prevention strategies from February 3^rd^ may have contributed to declines in population movement prior to the nationwide lockdown, particularly in March, when all mobile phone users received 10 SMS push notifications from the Ministry of Health in addition to information provided through other media; these early reductions in population movement may have contributed to lowering the reproduction number. The majority of imported cases were less than 30 years old (many were university students returning from studies abroad), and most of those that acquired the infection domestically were under 40 years, which may explain the low numbers with severe disease and absence of deaths.

The high proportion of cases that developed symptoms after isolation (73.9%) or never developed symptoms (43%) highlights one of the major challenges of controlling SARS-CoV-2 and the strengths of Vietnam’s approach. Suspected cases were identified and quarantined based on their epidemiological risk of infection (recent contact with a confirmed case or travel to a COVID-19 affected country), rather than on exhibiting symptoms. Without the implementation of strong control measures and meticulous contact-tracing, it is likely such cases would have silently transmitted the virus and undermined other control efforts.

The strength of our report is that it provides a complete picture based on national data of case numbers, their clinical and demographic characteristics, and the testing performed and various interventions made by the Government over time. Further, the use of systematic quarantine measures allowed clear distinction between imported and domestically-acquired cases, thus allowing for estimation of the efficiency of various interventions. The limitations are that the data are descriptive, contain relatively small numbers of confirmed cases, and only include the first 100 days of an epidemic that is likely to continue for many months. It is therefore impossible to conclude definitively which of these control measures have resulted in the current control of SARS-CoV-2 in Vietnam and whether they will continue to work in the future.

There are, however, two distinctive features of Vietnam’s response. First, the Government acted quickly, educating and engaging the public, placing restrictions on international flights, closing schools and universities, and instituting exhaustive case-contact tracing from late January, well before these measures were advised by WHO. Second, they placed the identification, serial testing, and minimum 14-day isolation of all direct contacts of cases, regardless of symptom development, at the heart of the response. Our findings suggest the latter measure was likely to be especially effective given nearly half of those infected did not develop symptoms.

In summary, Vietnam has controlled SARS-CoV-2 spread by acting early, maintaining clear and consistent public communications, introducing meticulous contact-tracing and quarantine, and implementing progressive international travel restrictions. The value of these interventions in controlling the infection is supported by the high proportion of asymptomatic cases and imported cases, and evidence for substantial pre-symptomatic transmission.

## Epilogue

At the time of submission (May 17^th^ 2020), lockdown measures have been progressively lifted and schools, universities and non-essential shops have been re-opened. Karaoke bars and places for mass gatherings remain closed. An additional 42 cases have been confirmed on arrival among repatriated Vietnamese nationals, and they have subsequently been isolated. No community transmission has been detected and no deaths have been recorded among the 312 cases, 52 of whom remain hospitalised / in isolation.

## Data Availability

Data available upon request

## Contributors

PQT, MAR, GT, HRvD, and MC conceived and designed the study, and wrote the manuscript. DHL, DQT, TDQ, and PCD provided the data for this study. TAT, HLQ, NAHT, TMP, VC, NDN, and NCK prepared the data. PQT, MAR, and MC performed the analyses. LNQ, DDA, and TND provided expertise on the study. All authors revised and approved the final version of the manuscript.

## Acknowledgements

We are grateful to the Ministry of Health of Vietnam and the National Steering Committee for COVID-19 response for making data available for this study, and Dr. Bui Vu Duy, the director of the Rapid Response Information Team of the National Steering Committee for COVID-19 Response, for his support. MR, GT, HRvD and MC and the OUCRU COVID-19 research group are supported by the Wellcome Trust.

## OUCRU COVID-19 Research Group Members

Mary Chambers, Marc Choisy, Jeremy Day, Dong Huu Khanh Trinh, Dong Thi Hoai Tam, Joseph Donovan, Du Hong Duc, Ronald B Geskus, Ho Quang Chanh, Ho Van Hien, Huong Dang Thao, Huynh le Anh Huy, Huynh Ngan Ha, Huynh Trung Trieu, Huynh Xuan Yen, Evelyne Kestelyn, Thomas Kesteman, Lam Anh Nguyet, Lam Minh Yen, Katrina Lawson, Le Kim Thanh, Le Nguyen Truc Nhu, Le Thanh Hoang Nhat, Le Thi Hoang Lan, Le Van Tan, Sonia Odette Lewycka, Nguyen Bao Tran, Nguyen Minh Nguyet, Nguyen Than Ha Quyen, Nguyen Thanh Ngoc, Nguyen Thi Han Ny, Nguyen Thi Hong Thuong, Nguyen Thi Huyen Trang, Nguyen Thi Kim Tuyen, Nguyen Thi Ngoc Diep, Nguyen Thi Phuong Dung, Nguyen Thi Tam, Nguyen Thi Thu Hong, Nguyen Thu Trang, Nguyen Van Vinh Chau, Nguyen Xuan Truong, Ninh Thi Thanh Van, Phan Nguyen Quoc Khanh, Phung Khanh Lam, Phung Le Kim Yen, Phung Tran Huy Nhat, Maia Rabaa, Thuong Nguyen Thuy Thuong, Guy Thwaites, Louise Thwaites, Tran My Phuc, Tran Tan Thanh, Tran Thi Bich Ngoc, Tran Tinh Hien, H Rogier van Doorn, Jennifer Van Nuil, Vinh Chau, Vu Thi Ngoc Bich, Vu Thi Ty Hang, Sophie Yacoub.

## 1 Data

### 1.1 Apple Maps Mobility Trends Reports

These data have been made available (https://www.apple.com/covid19/mobility) temporarily only, for the specific purpose of helping fighting against COVID-19^1^. Reports are published daily and reflect requests for directions in Apple Maps. This data is generated by counting the number of requests made to Apple Maps for directions, when moving by car or on foot. Data that is sent from users’ devices to the Maps service is associated with random, rotating identifiers so Apple doesn’t have a profile of the user’s movements and searches. The data show a relative volume of directions requests in Vietnam compared to a baseline volume on January 13^th^, 2020. Day are defined midnight-to-midnight, UTC-8. Relative volume varies from week to week, consistent with normal, seasonal usage of Apple Maps (e.g. the lunar New Year holiday). Day of week effects (week-end versus week days) are thus important to normalize when reading these data. Data that is sent from users’ devices to the Maps service is associated with random, rotating identifiers so Apple doesn’t have a profile of individual movements and searches. Apple Maps has no demographic information about our users, so we can’t make any statements about the representativeness of usage against the overall population.

### 1.2 Google Community Mobility Reports

These data have been made available (https://www.google.com/covid19/mobility) temporarily only, for the specific purpose of helping fighting against COVID-19^2^. These reports aim to provide insights into what has changed in response to policies aimed at combating COVID-19. The reports chart movement trends over time, across different categories of places such as **retail and recreation** (restaurants, cafes, shopping centers, theme parks, museums, libraries, and movie theaters), **groceries and pharmacies** (grocery markets, food warehouses, farmers markets, specialty food shops, drug stores, and pharmacies), **parks** (local parks, national parks, public beaches, marinas, dog parks, plazas, and public gardens), **transit stations** (public transport hubs such as subway, bus, and train stations), **workplaces**, and **residential**. No personally identifiable information, such as an individual’s location, contacts or movement, is made available at any point. Insights in these reports are created with aggregated, anonymized sets of data from users who have turned on the Location History setting, which is off by default. Artificial noise is also added in order to prevent the identification of any individual person. The baseline is the median value, for the corresponding day of the week, during the 5-week period Jan 3^rd^ - Feb 6^th^, 2020.

## 2 Methods

### 2.1 Serial intervals

A normal distribution was fitting by maximum likelihood to the values of serial intervals. The parameters (mean and standard deviation) estimates and their variance-covariance matrix were fed into a multinormal distribution in order to generate 10 000 Monte Carlo simulations of the serial interval distribution. We used these 10 000 simulated distributions to generate (i) the 95% confidence interval of the distribution of the serial intervals and (ii) the estimate, and its 95% confidence interval, of the proportion of distribution of the serial intervals that is below zero.

### 2.2 R packages used in the analysis

All analyses were performe in R 4.0.0^3^ with the following packages:

- fitdistrplus v. 1.0-14^4^ for fitting the normal distribution to serial intervals.
- mvtnorm v. 1.1-0^5^ for the multinormal distribution used in the Monte Carlo simulations.
- car v. 3.0-7^6^ for the Type-II analyses of deviance in order to correct for potential confounding effects.
- incidence v. 1.7.1^7^ to generate weekly incidence data from line listing. These weekly incidence data are then used to estimate weekly reproduction numbers.
- EpiEstim v. 2.2-1^8^ to estimate the reproduction numbers by week.

## 3. Results

An extended version of **Table 1** is shown in **Table S1**.

**Table S1.**
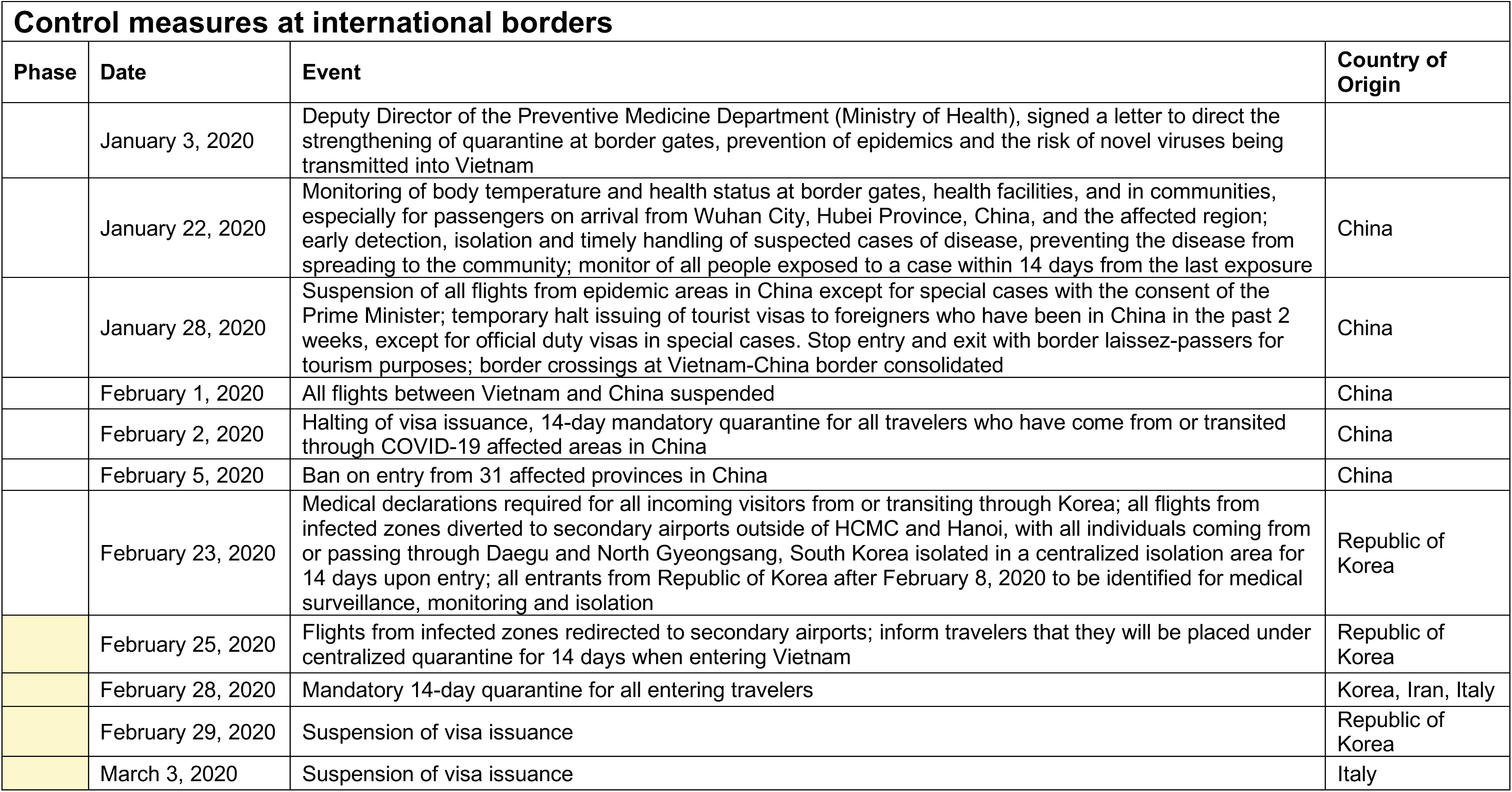
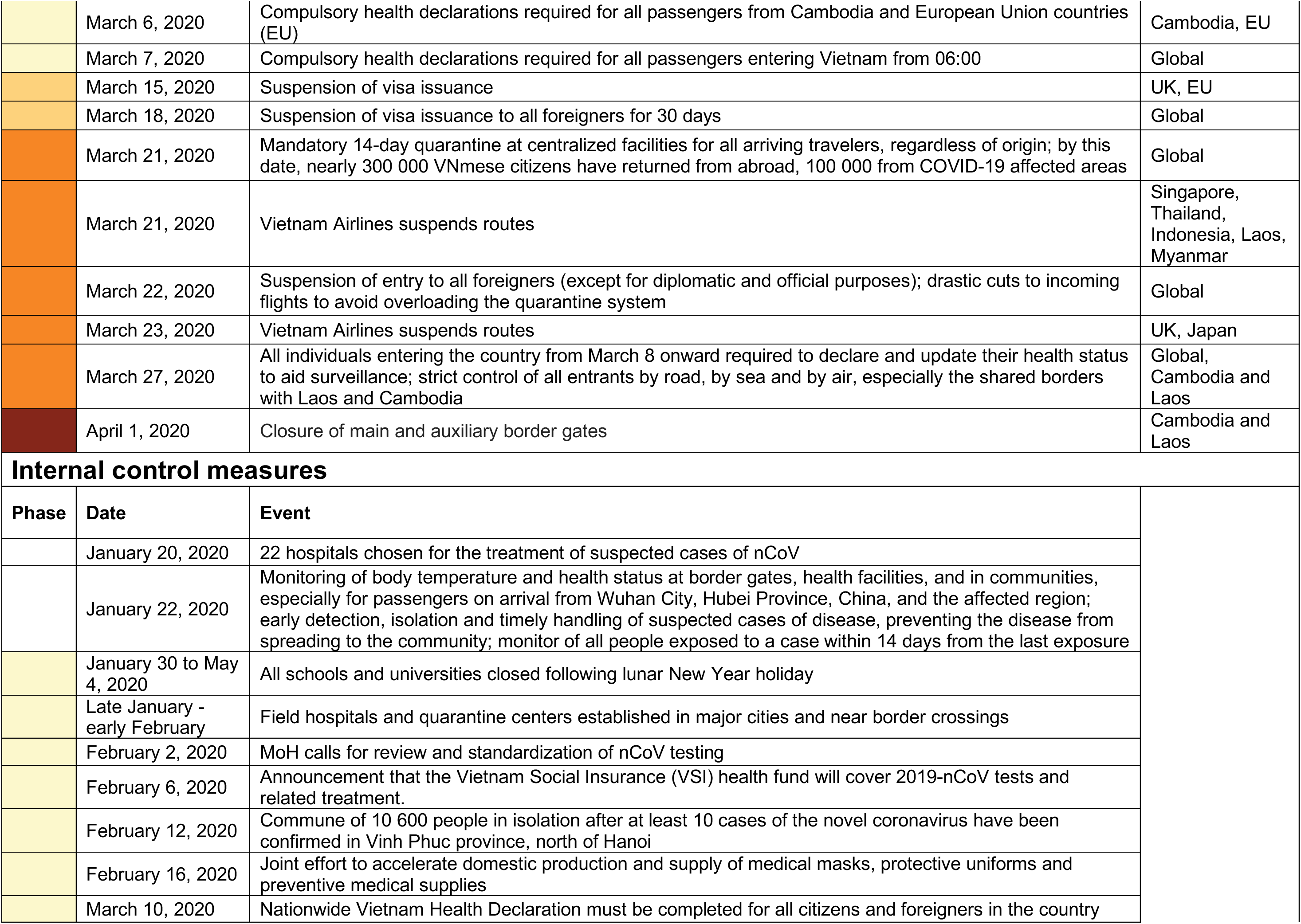
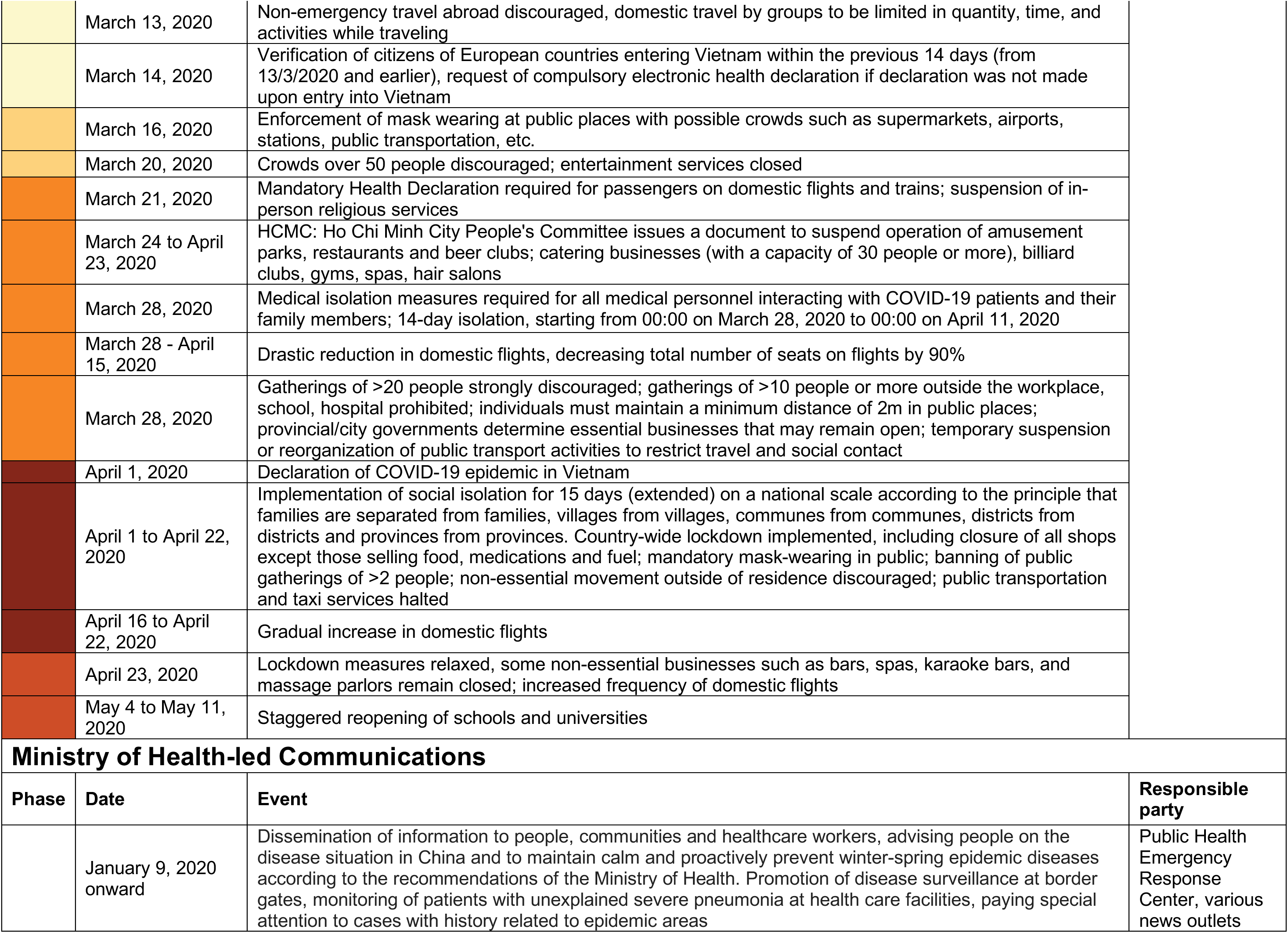
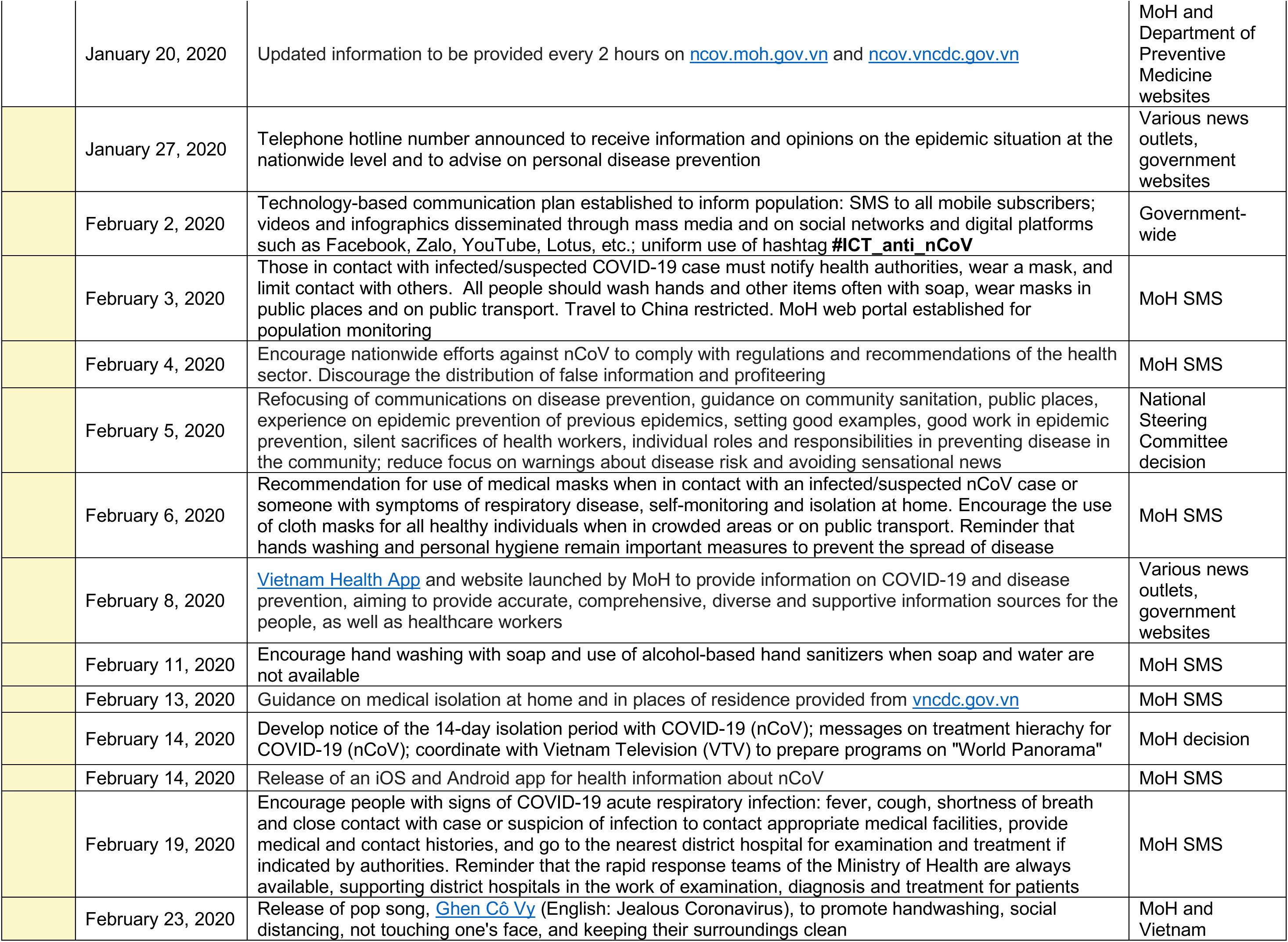
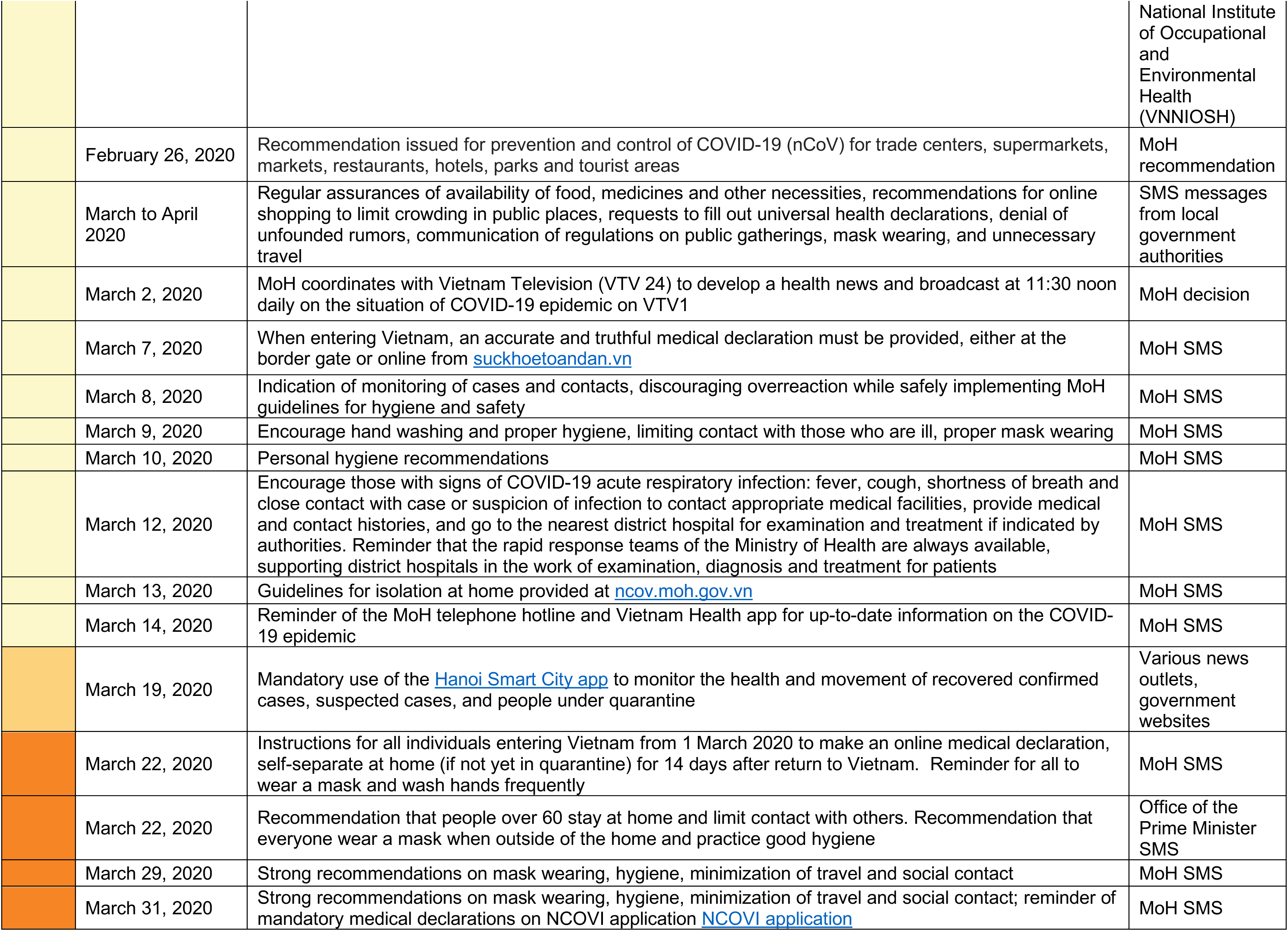
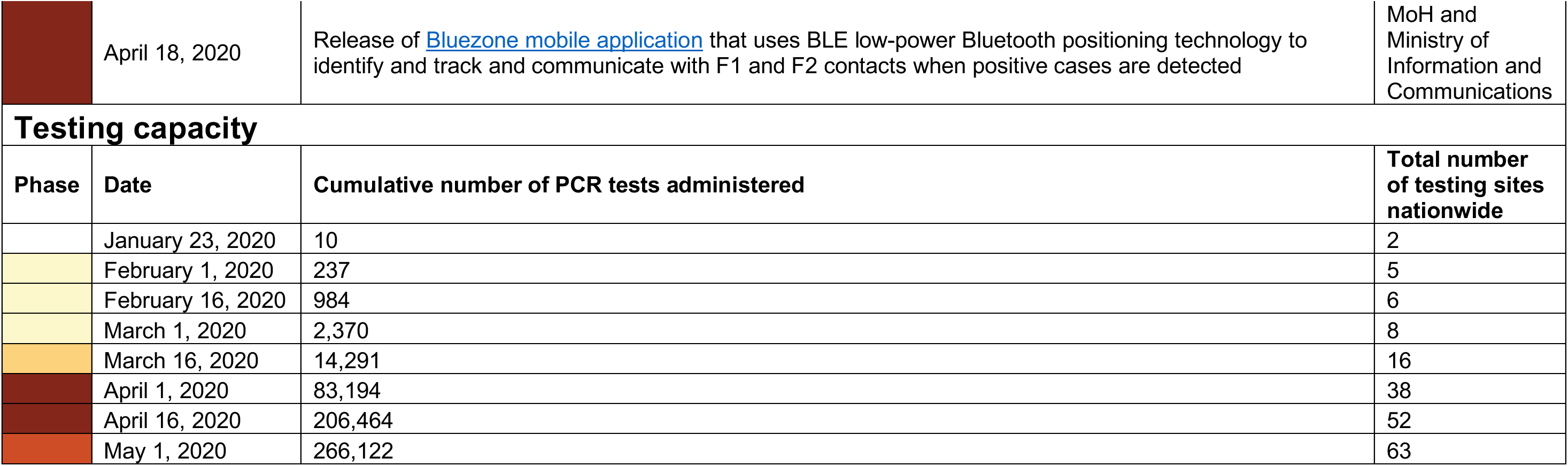
The timing and nature of major Government-led control measures, including international border control, internal control, Ministry of Health-led communications, and enhancement of diagnostic capability. The colors shown in the phase column indicate the intensity of control measures taken over different periods (white, initial; light yellow, early; light orange, intermediate; orange, pre-epidemic; brown, epidemic/lockdown; dark orange, post-lockdown), and correspond to the colors used in Table 1 and Figure 1 in the main text.

**Table S2.**
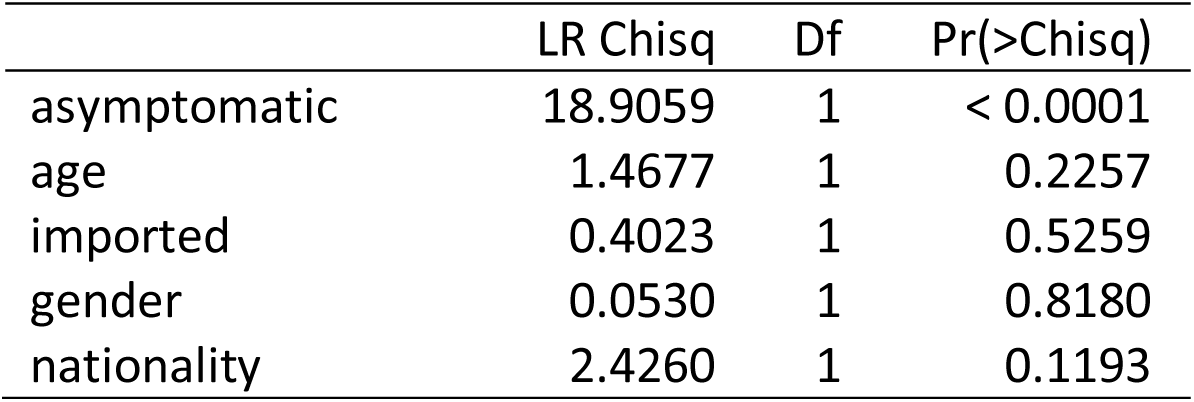
Analysis of deviance table (type-II tests) for Figure 3B explaining the duration of hospitalization with a gamma regression. LR Chisq: chi-squared statistic, Df: degrees of freedom, Pr(>Chisq): p-value of the type-II likelihood ratio test.

**Table S3.** Analysis of deviance table (type-II tests) for Figure 3C explaining the proportion asymptomatic with a logistic regression. LR Chisq: chi-squared statistic, Df: degrees of freedom, Pr(>Chisq): p-value of the type-II likelihood ratio test.

|  | LR Chisq | Df | Pr(>Chisq) |
| --- | --- | --- | --- |
| age | 0.1999 | 1 | 0.6548 |
| imported | 3.0757 | 1 | 0.0795 |
| gender | 0.0915 | 1 | 0.7623 |
| nationality | 0.3049 | 1 | 0.5808 |

